# Home monitoring of HbA1c in diabetes mellitus: A protocol for systematic review and narrative synthesis on reliability, accuracy, and patient acceptability

**DOI:** 10.1101/2021.12.15.21267851

**Authors:** Jack Colley, Hajira Dambha-Miller, Beth Stuart, Jazz Bartholomew, Hermione Price

**Affiliations:** Southern Health Foundation Trust; Primary Care Research Centre, University of Southampton

## Abstract

**Introduction:** Worldwide there are an estimated 463 million people with diabetes. [1] In the UK people with diabetes are offered an annual review including monitoring of Haemoglobin A1c (HbA1c). [2] [3] This can identify people with diabetes who are not meeting their glycaemic targets, enabling early intervention. Those who do not attend these reviews often have higher HbA1c levels and poorer health outcomes. [4] During the Coronavirus disease of 2019 (COVID-19) pandemic, there was a 77% reduction in monitoring of HbA1c in the UK. [5] We hypothesise that people with diabetes could take finger-prick samples at home for the measurement of HbA1c.

**Method and Analysis:** We will perform a systematic review of current evidence for capillary blood collected at home for the measurement of HbA1c. We will examine the validity, reliability, safety, and patient acceptability of the use of capillary blood compared with the usual standard of care of venous blood. We will explore variables which affect validity of results. Using core terms of ‘Diabetes’, ‘HbA1c’ and ‘Capillary sampling’ we will search MEDLINE, Embase, CINAHL, Web of Science Core Collection, Google Scholar, Open Grey and other grey literature from database inception until 2021. Risk of bias will be assessed using the ‘COSMIN risk of bias tool to assess the quality of studies on reliability and measurement error’. Database searches and data extraction for primary outcomes will be conducted in duplicate. We will produce a narrative synthesis exploring how variables of capillary blood collection impact on validity, as well as exploring the safety and acceptability of patient self-collection.

**Ethics and Dissemination:** This review will be submitted for publication in a peer-reviewed open-access journal. We will present our results at both national and international conferences. As a systematic review with no primary participant data or involvement, ethical approval is not applicable.

**PROSPERO registration number:** CRD42021225606

**Strengths and limitations of this study:**

- To our knowledge this is the first systematic review to explore all postal methods of capillary blood collection for the measurement of HbA1c.
- This review follows the Preferred Reporting Items for Systematic Reviews and Meta-Analyses guidelines, which offers transparency and enhances reproducibility.
- Due to anticipated heterogeneity in statistical approaches, summary analyses, sample storage, transportation, extraction, and assay, meta-analysis is unlikely to be appropriate and therefore narrative synthesis will be used.
- Due to the exclusion criteria, our findings may not be generalisable to a wider population including children, and those with haemoglobinopathies, high erythrocyte turnover or other conditions likely to affect HbA1c result.

## Background

Diabetes is one of the biggest health issues worldwide. Globally, there are an estimated 463 million people with the disease, a number that has increased four-fold over the last 35 years. [3] [6] [1] In the United Kingdom (UK), 4.9 million people are currently living with diabetes. [7] The prevalence of both type one and type two diabetes is increasing, however this rise is far more alarming in cases of type two diabetes. [8] [9]

The diabetes annual review process consists of a series of health checks recommended by the national service framework and the National Institute for Health and Care Excellence (NICE). [2] [10] These health checks include monitoring of HbA1c, blood pressure, cholesterol, kidney function, urinary albumin, Body Mass Index (BMI), and foot health. There is financial incentive for practices that record a high percentage of patients achieving recommended treatment goals for HbA1c, blood pressure and cholesterol. This process aims to improve standards of care by identifying where people with diabetes may not be meeting recommended targets, which can enable earlier intervention and prevent longer term morbidity.

During the COVID-19 pandemic, the introduction of social distancing led to many health care practitioners working remotely for delivery of routine care. People with diabetes were identified as clinically vulnerable and were advised to adhere stringently to social distancing guidance. [11] [12] [13] As a consequence, in April 2020 there was a reported 77% reduction in monitoring of HbA1c in England, as well as less recorded monitoring of blood pressure, cholesterol, creatinine, urinary albumin, and BMI. There was an estimated 60,000 missed or delayed new diagnoses of diabetes across the UK. [5]

Prior to the COVID-19 pandemic, non-attendance at diabetes outpatient appointments was a sizeable problem. In England in the 2019-20 financial year over six percent of all appointments were not attended, equating to over 7,000,000 appointments wasted, rates are similar in people with diabetes. [14] [15]Perhaps more concerningly a recent systematic review suggested non-attenders on average had higher HbA1c levels and worse health outcomes. [4] A system of required attendance at a healthcare setting in order to measure venous HbA1c will inevitably fail to fully address this cohort, who arguably are the most in need of diabetes surveillance.

We hypothesise that people with diabetes could take finger prick samples at home and post these to a laboratory to obtain an HbA1c result. There is a growing body of evidence surrounding the use of capillary blood and dried blood spots in particular for measuring HbA1c. [16] [17] [18] [19] [20] [21] [22] [23] However, there is still a need for standardisation of collection, storage and transportation methods. [16] [21] [23] To guide standardisation we need to know which methods can produce reliable and valid results, and which variables impact on bias and imprecision of results. We use validity as an all-encompassing term for reliability, precision, and accuracy. Accuracy is used to refer to the similarity of capillary and venous results from the same person. We define reliability as the ability to produce similar results from repeated samples taken over a clinically insignificant time frame. We define precision as whether a test can confidently give a HbA1c result to an exact number, or if less precise, whether it can still inform the clinician of good, moderate, or poor glycaemic control.

We also need to explore whether changes to current practice could introduce new risks, whether that be risks from the blood collection and transport procedure itself, or risks from reduced contact with health professionals as a result of self-collection at home. We also need to know if a self-collected HbA1c test would be acceptable to patients, and what might influence the uptake of such a test.

In this systematic review we aim to explore all currently available evidence examining HbA1c measured from capillary blood compared with venous results, and whether there are viable postal alternatives to venous sampling. We will examine which variables impact the validity of these results. The evidence produced could be a step towards transforming current diabetes practices, especially in more remote communities where the collection, storage and transport of venous blood samples may not be feasible. The COVID-19 pandemic has presented a clear and urgent need for improving access to healthcare remotely in people with diabetes.

## Objectives

- Are HbA1c levels determined from capillary blood obtained via finger-prick testing reliable, and are they valid when compared to venous blood sampling?
- Can HbA1c be measured safely at home by people with diabetes mellitus?
- Is self-collected finger-prick sampling at home, as a means of measuring HbA1c, acceptable to people with diabetes as an alternative to venous blood sampling in a healthcare setting?

## Study Criteria

### Population

Studies of adults aged 18 years or older who are being screened for, monitored, or previously diagnosed with diabetes mellitus. Studies which exclusively look at HbA1c measurement in people with haemoglobinopathies, anaemias, high erythrocyte turnover or other conditions known to affect HbA1c interpretation may be discussed but not included in analysis of validity. Animal studies will not be included.

### Inclusion criteria and Index test

Articles which examine methods of capillary blood self-collection and storage for measurement of HbA1c, which look at test reliability, validity, or patient acceptability as detailed above, will be included. We will not include studies of novel methods still in experimental stages, where validation studies with patient blood specimens have not been carried out. We will include studies using venous blood to test novel methods for capillary blood collection (e.g. the use of venous blood on a dried blood spot). These studies will be differentiated from capillary studies in our analysis. We will only include studies of methods which would be practical to post to people with diabetes, thus excluding most point of care methods or those requiring high intensity storage such as refrigeration. Studies which look at the effect of time, temperature, and other storage variables on capillary HbA1c results will be included.

### Target condition

Any form of diabetes mellitus where HbA1c would be used for diagnosis or monitoring. Routine use of HbA1c is not recommended in women in the second or third trimester of pregnancy and therefore studies in this cohort will not be included. [24]

### Reference standard

For studies of validity or reliability, the reference standard must be a venous blood test analysed by a certified method for HbA1c testing as listed by the International Federation of Clinical Chemistry and Laboratory Medicine (IFCC) or the National Glycohemoglobin Standardization Program (NGSP). [25] [26]We will exclude studies where the time interval between index test and reference standard is more than two weeks for >10% of participants.

## Methods

This protocol is reported according to the Preferred Reporting Items for Systematic Reviews and Meta-Analyses (PRISMA) guidance and has been registered on PROSPERO. [27] [28]

### Search strategy

The key search terms are ‘Diabetes’ AND ‘HbA1c’ AND ‘Capillary sampling’ which have been expanded in our full search strategy (Appendix 1 or available through PROSPERO (**registration number** CRD42021225606)). Free text, MESH terms, varied spellings and acronyms have been included to maximise capture of all relevant studies. We will search the Embase, MEDLINE, CINAHL and Web of Science Core Collection databases from database inception to September 2021. We will additionally search the grey literature through OpenGrey as well as using Google Scholar limiting retrieval to the first 200 relevant references as per guidance on optimal database combinations. [29] Ongoing studies and other grey literature will be sought from relevant conference abstracts from 2020 and 2021. Reference lists from the most relevant papers will be hand-searched using an ancestry approach. Results will be restricted to those available in the English language only.

### Study Selection and Data Management

Titles and abstracts from search results will be downloaded to Endnote X9, where they will be de-duplicated. They will then be uploaded to Rayyan.ai for review. Two reviewers (JC and JB) will independently screen titles and abstracts against inclusion and exclusion criteria whilst blinded to the other’s decisions. Selected studies will then undergo full text review by both reviewers. Any discrepancies that arise during this process will be discussed between the two reviewers until a resolution is reached. Where a resolution cannot be found a third reviewer will be consulted. A PRISMA flow diagram will be used to demonstrate study selection and exclusion. Studies which undergo full text review will be documented, and if excluded at this stage we will keep record of the reasons for exclusion and make this information available.

We have created a data extraction tool using Microsoft Excel (Version 2102) which was piloted (Appendix 2). Two reviewers will carry out data extraction (JC and JB) and disagreements resolved through discussion.

The data being collected will include information regarding:

- Study Characteristics (Such as country, year, setting, objectives of the study, study inclusion and exclusion criteria)
- Participant Characteristics (Such as population description, age, sex, total number, diabetic status, and dropout rates)
- The capillary blood method/methods being investigated (including details about method used, person collecting the sample, location, storage and shipping conditions, laboratory methods including analysis device, laboratory blinding, resource requirements and timing between test and reference)
- The reference standard (including analysis device used, location of collection, storage and shipping conditions, laboratory blinding and number of available results)
- The primary outcome (including statistical methods used and outcomes relevant to that method, diagnostic thresholds if used, measures of error and bias and any other reported results)
- Additional outcomes (Patient acceptability, and where evidence exists to do so we will collect information on facilitators and barriers to patient uptake)

### Quality assessment

We will be using ‘COnsensus-based Standards for the selection of health Measurement INstruments (COSMIN) risk of bias tool to assess the quality of studies on reliability and measurement’ to assess the quality of selected studies. The COSMIN tool will be piloted by both reviewers (JC and JB) on the first five selected studies. Following this if there is good agreement between both reviewers then the quality assessment for the remaining studies will be carried out by one reviewer only (JC). If there is poor agreement during the pilot this will be discussed between the two reviewers, the tool will be revised or replaced, repiloted, and this process will continue until good agreement is reached. As we are performing narrative synthesis, there will be no limit on what level of quality leads to inclusion. Instead, we will discuss quality of studies as part of the synthesis of their results.

### Data Synthesis

From preliminary searches we have identified methodological heterogeneity in the studies identified. There is variability in how data has been presented, studies have either presented outcomes in terms of sensitivity and specificity of HbA1c as a diagnostic test, or at pre-defined clinically significant levels, or as measures of error, with correlation co-efficients, intercepts, Bland-Altman plots, and error grid analyses. There is also heterogeneity in the collection, storage and transportation methods and type of blood used for the index test. Full exploration of heterogeneity will take place following completion of searches, by visual examination of tables ordered by likely modifiers. Identified modifiers will be used to determine subgroups for synthesis, which will in turn be used to determine how these variables can impact validity.

Given the predicted level of heterogeneity, we anticipate the most appropriate method of synthesis will be narrative. We will be following the Synthesis Without Meta-Analysis (SWiM) reporting guidelines adapted for a non-interventional review. [30] The SWIM guidance is tailored towards reviews of intervention, and therefore it may need adapting as we undertake our synthesis. Where significant heterogeneity exists, meta-analysis can often fail to produce meaningful results by under-representing the overall body of evidence. Narrative synthesis will enable us to incorporate all available evidence despite heterogeneity.

We will discuss measurement outcomes and the appropriateness of each statistical method used to the study design. We will summarise outcomes by their direction of effect (which we define as whether the index test is or isn’t valid or reliable for measurement of HbA1c compared to reference standard), and use vote counting to inform our conclusions. We hope to not only present the current evidence, but also identify key gaps in the literature to guide future research.

For the secondary outcome of patient acceptability, the method of synthesis will depend largely on the amount of data available. We would ideally want to perform qualitative synthesis through the use of thematic analysis to further understand facilitators and barriers to patient uptake of home HbA1c measurement. If evidence for patient acceptability is limited, we will instead look at uptake and dropout rates, using this information to infer probable popularity of a proposed at home collection option.

### Grading the strength of recommendations

We will be using the Grading of Recommendations, Assessment, Development and Evaluations framework (GRADE) in making our recommendations. [31]

### Ethics and Dissemination

As a secondary analysis of existing published data with no primary identifiable personal data being collected no ethics review was required.

This review will be submitted for publication in a peer-reviewed open-access journal. Where invited to do so we will present our results at both national and international relevant conferences. These methods will allow dissemination to patient groups, clinicians, and guideline developers.

### Patient and Public Involvement

Patients or the public were not involved in the design of this review.

## Supporting information

Appendix 1: Search Strategy

Appendix 2: Data Extraction Tool

## Data Availability

All data produced in the present study are available upon reasonable request to the authors

## Author Statement

JC and HP were involved in the conceptualisation of the study. JC, HP, HDM and JB made substantial contributions to the design and development of the methodology. BS made substantial contributions to the statistical analysis methodology. JC was responsible for drafting the protocol manuscript. All authors reviewed, edited, and approved the final manuscript.

## Appendices

### Appendix 1

#### Search Strategies

##### Medline Search Strategy

1. exp Diabetes Mellitus/
2. Diabet*.mp.
3. T1DM.mp.
4. T2DM.mp.
5. “Impaired glucose tolerance”.mp.
6. “Impaired fasting glucose”.mp.
7. pre-diabetes.mp.
8. prediabetes.mp.
9. LADA.mp.
10. MODY.mp.
11. IDDM.mp.
12. NIDDM.mp.
13. Hyperglyc*.mp.
14. 1 or 2 or 3 or 4 or 5 or 6 or 7 or 8 or 9 or 10 or 11 or 12 or 13
15. Glycated Hemoglobin A/
16. HbA1c.mp.
17. A1c.mp.
18. HgbA1c.mp.
19. Hb1c.mp.
20. “Hb A1”.mp.
21. “Hb A1a+b”.mp.
22. “Hb A1b”.mp.
23. HbA1.mp.
24. “Glycated H?emoglobin”.mp.
25. “Glycosylated h?emoglobin”.mp.
26. Glycoh?emoglobin.mp.
27. “H?emoglobin A(1)”.mp.
28. “H?emoglobin A, Glycosylated”.mp.
29. “H?emoglobin, glycosylated”.mp.
30. 15 or 16 or 17 or 18 or 19 or 20 or 21 or 22 or 23 or 24 or 25 or 26 or 27 or 28 or 29
31. “H?emoglobin A1c Test Kit”.mp.
32. “capillary blood sampling”.mp.
33. “finger prick”.mp.
34. fingerprick.mp.
35. “finger stick”.mp.
36. fingerstick.mp.
37. “Heel prick”.mp.
38. heelprick.mp.
39. Lancet.mp.
40. Lancing.mp.
41. Dried Blood Spot Testing/
42. “Dry blood spot”.mp.
43. DBS.mp.
44. “capillary tube”.mp.
45. microtube.mp.
46. ”micro tube”.mp.
47. “P?ediatric tube”.mp.
48. “blood sampling device”.mp.
49. “blood sampling needle”.mp.
50. Point-of-Care Systems/
51. point-of-care systems/ or point-of-care testing/
52. “point of care”.mp.
53. “blood collection paper”.mp.
54. 31 or 32 or 33 or 34 or 35 or 36 or 37 or 38 or 39 or 40 or 41 or 42 or 43 or 44 or 45 or 46 or 47 or 48 or 49 or 50 or 51 or 52 or 53
55. 14 and 30 and 54
56. limit 55 to (english language and humans)

##### Embase Search Strategy

1. exp diabetes mellitus/
2. diabet*.mp.
3. “Impaired fasting glucose”.mp.
4. pre?diabetes.mp.
5. LADA.mp.
6. MODY.mp.
7. hyperglycemia/
8. 1 or 2 or 3 or 4 or 5 or 6 or 7
9. exp hemoglobin A1c/
10. A1c.mp.
11. HgbA1c.mp.
12. Hb1c.mp.
13. “Hb A1”.mp.
14. “Hb A1a+b”.mp.
15. “Hb A1b”.mp.
16. HbA1.mp.
17. “Glycosylated Hemoglobin”.mp.
18. “Hemoglobin A(1)”.mp.
19. “Haemoglobin A(1)”.mp.
20. “HbA(1c)”.mp.
21. 9 or 10 or 11 or 12 or 13 or 14 or 15 or 16 or 17 or 18 or 19 or 20
22. dried blood spot testing/
23. exp hemoglobin A1c test kit/
24. capillary blood/
25. “capillary blood sampling”.mp.
26. “Finger prick”.mp.
27. Fingerprick.mp.
28. “Finger stick”.mp.
29. Fingerstick.mp.
30. DBS.mp.
31. “Heel prick”.mp.
32. Heelprick.mp.
33. Lancet.mp.
34. capillary tube/
35. microtube.mp.
36. “micro tube”.mp.
37. “P?ediatric tube”.mp.
38. “Lancing device”.mp.
39. blood sampling device/
40. blood collection needle/
41. ”blood collection paper”.mp.
42. “point of care system”/ or “point of care testing”/
43. 22 or 23 or 24 or 25 or 26 or 27 or 28 or 29 or 30 or 31 or 32 or 33 or 34 or 35 or 36 or 37 or 38 or 39 or 40 or 41 or 42
44. 8 and 21 and 43
45. limit 44 to (human and english language)

##### CINAHL search strategy

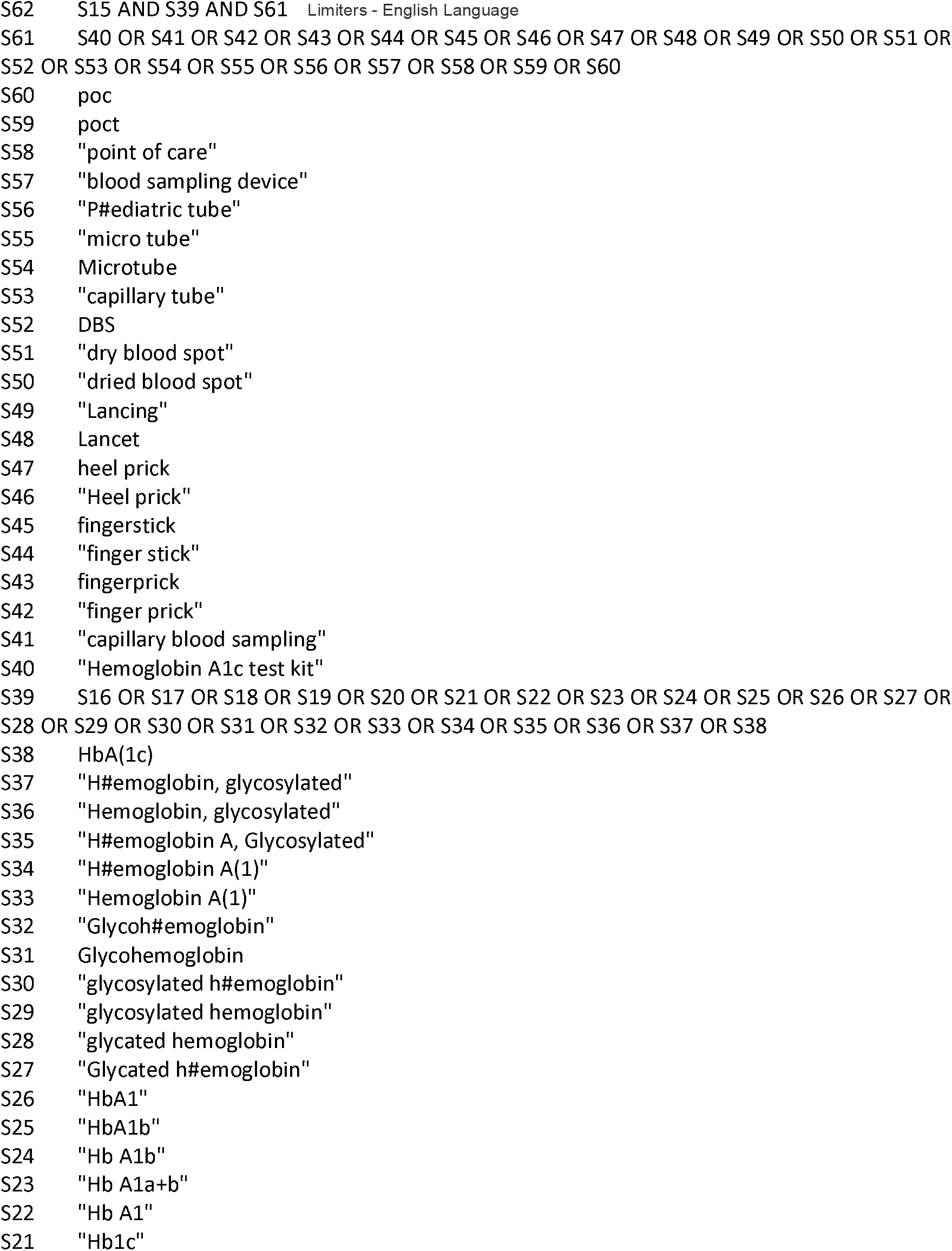
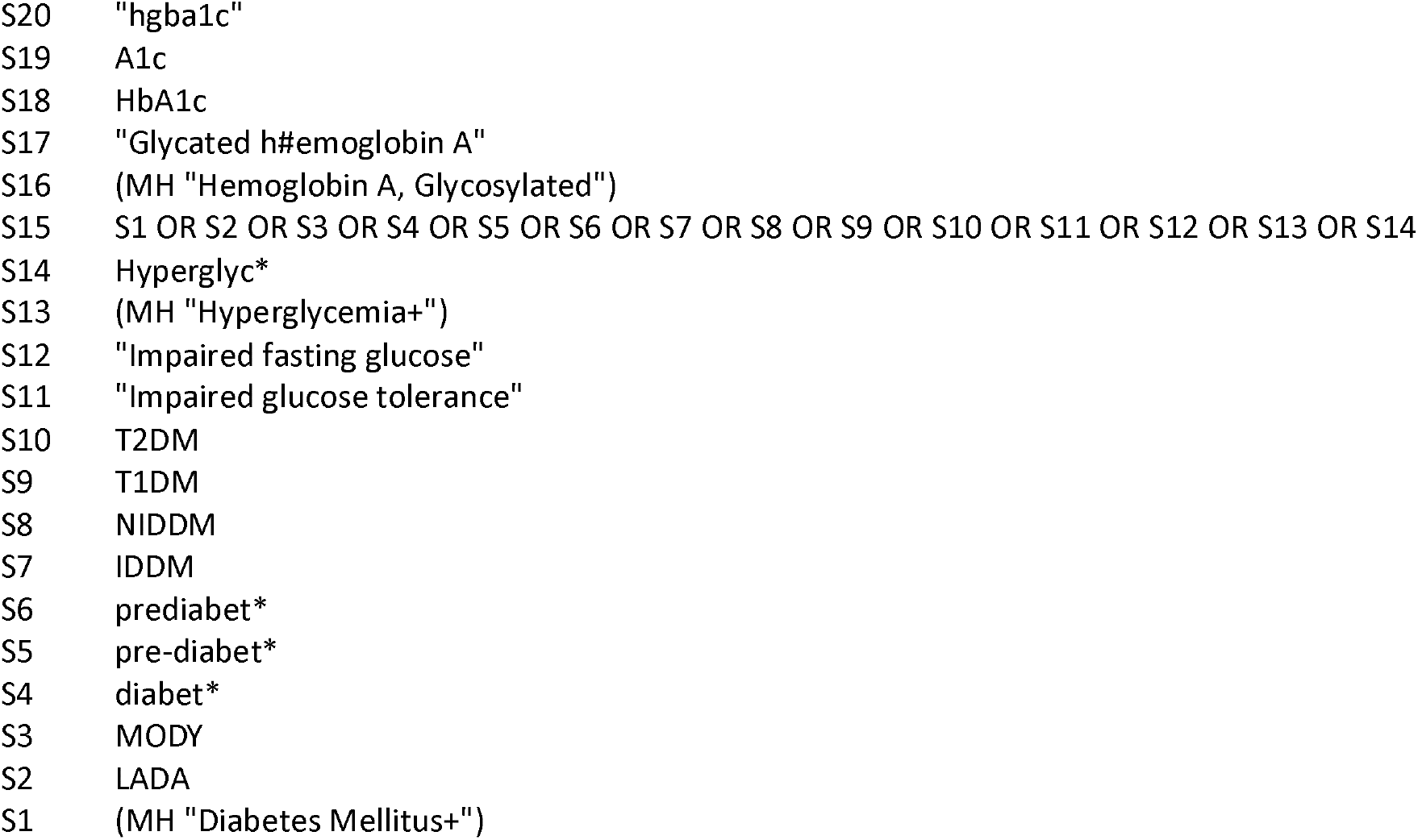

##### Web of Science: Core Collection search strategy

50 ((#13) AND #27) AND #48 and English (Languages)

49 ((#13) AND #27) AND #48

48 (((((((((((((((((((#28) OR #29) OR #30) OR #31) OR #32) OR #33) OR #34) OR #35) OR #36) OR #37) OR #38) OR #39) OR #40) OR #41) OR #42) OR #43) OR #44) OR #45) OR #46) OR #47

47 ALL=(POCT)

46 ALL=(POC)

45 ALL=(“Point of care”)

44 ALL=(“Blood sampling”)

43 ALL=(“P*ediatric tube”)

42 ALL=(“micro tube”)

41 ALL=(Microtube)

40 ALL=(DBS)

39 ALL=(“Dry Blood Spot”)

38 ALL=(“Dried Blood Spot”)

37 ALL=(“Capillary tube”)

36 ALL=(Lancing)

35 ALL=(Lancet)

34 ALL=(“Heel prick”)

33 ALL=(Heelprick)

32 ALL=(“Finger stick”)

31 ALL=(Fingerstick)

30 ALL=(“finger prick”)

29 ALL=(Fingerprick)

28 ALL=(“Capillary blood”)

27 ((((((((((((#14) OR #15) OR #16) OR #17) OR #18) OR #19) OR #20) OR #21) OR #22) OR #23) OR #24) OR #25) OR #26

26 ALL=(HbA(1c))

25 ALL=(“H*emoglobin, glycosylated”)

24 ALL=(“H*emoglobin A, Glycosylated”)

23 ALL=(“H*emoglobin A(1)”)

22 ALL=(glycoh*emoglobin)

21 ALL=(“glycosylated h*emoglobin”)

20 ALL=(“Glycated h*emoglobin”)

19 ALL=(HbA1)

18 ALL=(“Hb A1”)

17 ALL=(Hb1c)

16 ALL=(HgbA1c)

15 ALL=(A1c)

14 ALL=(HbA1c)

13 (((((((((((#1) OR #2) OR #3) OR #4) OR #5) OR #6) OR #7) OR #8) OR #9) OR #10) OR #11) OR #12

12 ALL=(Hyperglyc*)

11 ALL=(NIDDM)

10 ALL=(IDDM)

9 ALL=(MODY)

8 ALL=(LADA)

7 ALL=(“Impaired fasting glucose”)

6 ALL=(“Impaired glucose tolerance”)

5 ALL=(T2DM)

4 ALL=(T1DM)

3 ALL=(pre-diabet*)

2 ALL=(prediabet*)

1 ALL=(Diabet*)

##### Google Scholar search strategy

Diabetes|Diabetic|T1DM|T2DM|MODY|IDDM|NIDDM|Hyperglycaemia HbA1c|A1c|”Glycated haemoglobin”|”Glycated haemoglobin”|”Glycosylated haemoglobin”|”Glycosylated haemoglobin” “Finger prick”|”Hemoglobin A1c test kit”|DBS|”Dried blood spot”|Capillary|Lancet

