## Appendix 1: Search Strategy for "Home monitoring of HbA1c in diabetes mellitus: A protocol for systematic review and narrative synthesis on reliability, accuracy, and patient acceptability"

S62	S15 AND S39 AND S61	Limiters - English Language

S61	S40 OR S41 OR S42 OR S43 OR S44 OR S45 OR S46 OR S47 OR S48 OR S49 OR S50 OR S51 OR S52 OR S53 OR S54 OR S55 OR S56 OR S57 OR S58 OR S59 OR S60

S60	poc

S59	poct

S58	"point of care"

S57	"blood sampling device"

S56	"P#ediatric tube"

S55	"micro tube"

S54	Microtube

S53	"capillary tube"

S52	DBS

S51	"dry blood spot"

S50	"dried blood spot"

S49	"Lancing"

S48	Lancet

S47	heel prick

S46	"Heel prick"

S45	fingerstick

S44	"finger stick"

S43	fingerprick

S42	"finger prick"

S41	"capillary blood sampling"

S40	"Hemoglobin A1c test kit"

S39	S16 OR S17 OR S18 OR S19 OR S20 OR S21 OR S22 OR S23 OR S24 OR S25 OR S26 OR S27 OR S28 OR S29 OR S30 OR S31 OR S32 OR S33 OR S34 OR S35 OR S36 OR S37 OR S38

S38	HbA(1c)

S37	"H#emoglobin, glycosylated"

S36	"Hemoglobin, glycosylated"

S35	"H#emoglobin A, Glycosylated"

S34	"H#emoglobin A(1)"

S33	"Hemoglobin A(1)"

S32	"Glycoh#emoglobin"

S31	Glycohemoglobin

S30	"glycosylated h#emoglobin"

S29	"glycosylated hemoglobin"

S28	"glycated hemoglobin"

S27	"Glycated h#emoglobin"

S26	"HbA1"

S25	"HbA1b"

S24	"Hb A1b"

S23	"Hb A1a+b"

S22	"Hb A1"

S21	"Hb1c"

S20	"hgba1c"

S19	A1c

S18	HbA1c

S17	"Glycated h#emoglobin A"

S16	(MH "Hemoglobin A, Glycosylated")

S15	S1 OR S2 OR S3 OR S4 OR S5 OR S6 OR S7 OR S8 OR S9 OR S10 OR S11 OR S12 OR S13 OR S14

S14	Hyperglyc*

S13	(MH "Hyperglycemia+")

S12	"Impaired fasting glucose"

S11	"Impaired glucose tolerance"

S10	T2DM

S9	T1DM

S8	NIDDM

S7	IDDM

S6	prediabet*

S5	pre-diabet*

S4	diabet*

S3	MODY

S2	LADA

S1	(MH "Diabetes Mellitus+")

**Web of Science: Core Collection search strategy**

50 ((#13) AND #27) AND #48 and English (Languages)

49 ((#13) AND #27) AND #48

48 (((((((((((((((((((#28) OR #29) OR #30) OR #31) OR #32) OR #33) OR #34) OR #35) OR #36) OR #37) OR #38) OR #39) OR #40) OR #41) OR #42) OR #43) OR #44) OR #45) OR #46) OR #47

47 ALL=(POCT)

46 ALL=(POC)

45 ALL=("Point of care")

44 ALL=("Blood sampling")

43 ALL=("P*ediatric tube")

42 ALL=("micro tube")

41 ALL=(Microtube)

40 ALL=(DBS)

39 ALL=("Dry Blood Spot")

38 ALL=("Dried Blood Spot")

37 ALL=("Capillary tube")

36 ALL=(Lancing)

35 ALL=(Lancet)

34 ALL=("Heel prick")

33 ALL=(Heelprick)

32 ALL=("Finger stick")

31 ALL=(Fingerstick)

30 ALL=("finger prick")

29 ALL=(Fingerprick)

28 ALL=("Capillary blood")

27 ((((((((((((#14) OR #15) OR #16) OR #17) OR #18) OR #19) OR #20) OR #21) OR #22) OR #23) OR #24) OR #25) OR #26

26 ALL=(HbA(1c))

25 ALL=("H*emoglobin, glycosylated")

24 ALL=("H*emoglobin A, Glycosylated")

23 ALL=("H*emoglobin A(1)")

22 ALL=(glycoh*emoglobin)

21 ALL=("glycosylated h*emoglobin")

20 ALL=("Glycated h*emoglobin")

19 ALL=(HbA1)

18 ALL=("Hb A1")

17 ALL=(Hb1c)

16 ALL=(HgbA1c)

15 ALL=(A1c)

14 ALL=(HbA1c)

13 (((((((((((#1) OR #2) OR #3) OR #4) OR #5) OR #6) OR #7) OR #8) OR #9) OR #10) OR #11) OR #12

12 ALL=(Hyperglyc*)

11 ALL=(NIDDM)

10 ALL=(IDDM)

9 ALL=(MODY)

8 ALL=(LADA)

7 ALL=("Impaired fasting glucose")

6 ALL=("Impaired glucose tolerance")

5 ALL=(T2DM)

4 ALL=(T1DM)

3 ALL=(pre-diabet*)

2 ALL=(prediabet*)

1 ALL=(Diabet*)

**Google Scholar search strategy**

Diabetes|Diabetic|T1DM|T2DM|MODY|IDDM|NIDDM|Hyperglycaemia HbA1c|A1c|”Glycated haemoglobin”|”Glycated haemoglobin”|”Glycosylated haemoglobin”|”Glycosylated haemoglobin” “Finger prick”|”Hemoglobin A1c test kit”|DBS|”Dried blood spot”|Capillary|Lancet
